# Relationship Between Kinesiophobia and Fear of Falling in Patients Suffering from Stroke Leading to Physical Disability in Selected Rehabilitation Center of Bangladesh

**DOI:** 10.1101/2025.08.01.25332781

**Authors:** Arpon Kumar Paul, Polok Halder, Md. Abdul Alim

## Abstract

**Background:** Psychological barriers in stroke rehabilitation remain understudied in low-resource settings. This cross-sectional study examines relationships between kinesiophobia, fear of falling (FOF), and physical disability in Bangladeshi stroke patients.

**Methods:** Using validated Bangla versions of WHODAS 2.0 (12-item), Tampa Scale for Kinesiophobia (TSK-17), and Falls Efficacy Scale-International (FES-I), we assessed 200 patients from two rehabilitation centers. Pearson correlations and linear regression analyzed associations between psychological factors and disability.

**Results:** Participants showed moderate-severe scores: WHODAS 2.0 (M = 43.65 ±6.78), TSK (M = 47.26 ±5.44), FES-I (M = 49.12 ±7.20). Strong correlations emerged between WHODAS-FES-I (r = 0.65, 95% CI: 0.58–0.71) and TSK-FES-I (r = 0.55, 95% CI: 0.47–0.62). Regression models identified age ≥56 (β = 0.34, p = 0.002) and female gender (β = 0.28, p = 0.008) as significant predictors of higher psychological scores.

**Conclusion:** Psychological factors strongly correlate with physical disability in Bangladeshi stroke survivors, with demographic predictors suggesting the need for gender- and age-specific interventions. Study limitations include recruitment from two urban centers and a cross-sectional design. Integration of psychological assessment in rehabilitation protocols is recommended.

## INTRODUCTION

Stroke is a leading cause of death and long-term disability worldwide, with profound social and economic consequences [1]. In Bangladesh, the incidence of stroke is rising alarmingly due to factors such as urbanization, poor diet, and inadequate physical activity, making it a major public health challenge [1]. Stroke survivors often experience not only physical impairments like hemiplegia and postural instability but also significant psychological barriers that impede their recovery. Among these, fear of falling (FOF) and kinesiophobia—fear of movement due to the perceived risk of re-injury or pain—are increasingly recognized as critical factors that limit mobility, independence, and overall quality of life.

The purpose of this article is to investigate the relationship between kinesiophobia and FOF in stroke patients undergoing rehabilitation in Bangladesh and to assess how these psychological factors correlate with physical disability. The rationale for this study is rooted in the observation that, despite the high prevalence of stroke-related disability in Bangladesh, the psychological aspects of recovery, particularly kinesiophobia and FOF, remain understudied and inadequately addressed in current rehabilitation protocols [1-3]. Most existing research and clinical services focus on physical rehabilitation, often neglecting the emotional and cognitive barriers that can hinder progress [4, 5].

Previous studies have established that both kinesiophobia and FOF are prevalent among stroke survivors and are associated with avoidance behaviors, reduced physical activity, muscle weakness, and increased risk of falls [3, 5, 6]. These factors contribute to a cycle of deconditioning and further disability, making rehabilitation efforts less effective. In Bangladesh, where access to specialized rehabilitation services is limited, especially in rural areas, the impact of these psychological barriers is likely to be even more pronounced [4]. The lack of research on the interplay between kinesiophobia, FOF, and physical disability in this context underscores the need for targeted studies to inform more comprehensive and effective rehabilitation strategies.

Furthermore, studies in similar settings have highlighted the importance of addressing psychological factors such as anxiety, depression, and fear-related avoidance in stroke rehabilitation [2, 3, 6]. These findings suggest that integrating psychological support with physical therapy could improve rehabilitation outcomes. However, the specific relationship between kinesiophobia, FOF, and disability among Bangladeshi stroke patients has not been thoroughly investigated. This gap in knowledge is particularly significant given the country’s high stroke burden and limited rehabilitation resources [1, 4].

Therefore, this study aims to quantify the levels of kinesiophobia and FOF among stroke patients in Bangladesh, explore their association with physical disability, and identify demographic factors that may influence these outcomes. By doing so, it seeks to provide evidence that can guide the development of more holistic rehabilitation programs that address both the physical and psychological needs of stroke survivors. Ultimately, understanding and addressing these psychological barriers is essential for improving functional independence, quality of life, and long-term recovery in this vulnerable population.

## METHODS

### Study design

This study was conducted using a cross-sectional survey under a quantitative study design. Survey methodology was chosen to meet the study aim as an effective way to collect data. The recruitment period for this study extended from 22/02/2022 to 25/08/2022, conducted at the Centre for the Rehabilitation of the Paralysed (CRP) in Savar and the Centre for the Rehabilitation of the Paralysed (CRP) in Mirpur.

### Study settings

This study was conducted at two prominent rehabilitation centers in Bangladesh: The Centre for the Rehabilitation of the Paralysed (CRP) in Savar and the Centre for the Rehabilitation of the Paralysed (CRP) in Mirpur. These facilities are among the leading institutions providing specialized stroke rehabilitation services in the country, catering to a diverse population of stroke survivors from both urban and rural areas. The study here used a convenience sampling technique, considering the inclusion and exclusion criteria.

### Study population

Peoples who were suffering from stroke leading to physical disability was collected using convenience sampling from tertiary-level rehabilitation hospitals like the Centre for the Rehabilitation of the Paralyzed (CRP).

### Sample size calculation

A sample refers to a group of subjects selected from a population for research purposes [7]. In this study, although the ideal sample size was calculated as 384 using the finite population correction formula for a cross-sectional study—where Z = 1.96 (95% confidence interval), p = 0.5 (prevalence), q = 1 - p = 0.5, and d = 0.05 (margin of error)—only 200 stroke patients were included because of limitations in time and resources.

### Eligibility criteria

To determine if participants meet the inclusion and exclusion criteria for the study, a screening process was conducted.

#### Inclusion criteria

To be eligible to participate, participants needed to fulfill the following requirements: 1) Patient is being diagnosed stroke by MDT team, 2) All type of stroke (ischemic and hemorrhagic), 3) Male and female both are included, 4) Voluntary participation, 5) Patients having or done MRI or CT scan of head, and 6) First conducting patients.

#### Exclusion criteria

Participants were excluded if they had: 1) Patients having head injury or fracture, 2) Psychologically unstable patients, 3) Patients having brain tumour or malignancy, and 4) Patients who are not interested.

### Outcome measurement tools

#### Disability related questionnaire (WHODAS 12-item version)

The WHODAS 2.0 was developed to assess difficulties due to health conditions, including diseases, illnesses or injuries, mental or emotional problems, and problems with alcohol or drugs. The WHODAS 2.0 does not attempt to determine whether disability is due to physical or psychological disorders. The WHODAS 2.0 (like other generic disability measures that are not disorder-specific) has generally found the disability associated with mental disorders to be equal to or greater than that associated with physical disorders, depending on the specific mental and physical disorders being compared [8].

#### Kinesiophobia-related questionnaire (Tampa scale for Kinesiophobia)

The TSK is a 17-item self-report checklist using a 4-point Likert scale that was developed as a measure of fear of movement or (re)injury. Kinesiophobia is defined by the developers as “an irrational, and debilitating fear of physical movement and activity resulting from a feeling of vulnerability to painful injury or re-injury.” [9]. The scale is based on the model of fear avoidance, fear of work-related activities, fear of movement, and fear of re-injury [10]. The TSK has also been linked to elements of catastrophic thinking [11]. The scale can be useful in measuring unhelpful thoughts and beliefs about pain in people with chronic pain or fibromyalgia.

#### Fear of falling-related questionnaire (Falls Efficacy Scale)

The Fall Efficacy Scale-International (FES-I) is a questionnaire that assesses fear of falling (FOF) [12]. Fear of falling has been defined as an ongoing concern about falling, which ultimately limits the performance of activities of daily living [13]. Individuals are asked to rate, on a four-point Likert scale, their concerns about the possibility of falling when performing 16 activities. Individuals are instructed to rate each activity regardless of whether they actually perform it. The scores are added up to calculate a total score that ranges from 16 to 64 for the FES-I [14]. A higher score indicates a greater FOF.

### Study procedure and data collection methods

The researcher ensured ethical compliance by informing participants of their right to refuse any question and withdraw at any time. The study’s purpose was clearly communicated, and written consent was obtained before data collection. Participants were assured their personal information would remain confidential. Face-to-face interviews were conducted using a standardized Bangla-format questionnaire, allowing the researcher to clarify questions and maintain focus by minimizing environmental distractions. Interviews were conducted individually whenever possible to avoid external influence on responses. All data were collected directly by the researcher to reduce errors. The data analysis followed descriptive statistical methods using SPSS version 20.0, where variables were systematically labeled and coded. The dataset was cleaned to ensure accuracy before analysis. Frequency distributions, contingency tables, means, standard deviations, and chi-square tests were used to describe and explore associations between variables. Results were presented using tables, bar graphs, and pie charts, designed with Microsoft Excel 2007. The study followed the STROBE guidelines to maintain transparency and reliability in reporting cross-sectional observational research.

### Informed Consent

Verbal and written informed consent were obtained from every patient. Every patient was assured that they could leave at any time during data collection, and it was ensured that participants were not influenced by the data collector. The researcher strictly maintained confidentiality regarding participants’ conditions and treatments. The study was conducted in a clean and systematic manner. Every subject had the opportunity to discuss their problems with the senior authority or administration of CRP and had any questions answered to their satisfaction.

### Ethical considerations

Ethical considerations were strictly maintained in all aspects of the study, as they are a crucial component of all forms of research. The assessment files were securely stored and were not accessible to anyone other than the researcher. Written consent (Appendix) was obtained from all participants prior to completing the questionnaire. The researcher thoroughly explained their role in the study to the participants. Each participant provided a signed informed consent form, confirming their understanding of the study and their voluntary participation. Participants were clearly informed that their information would be kept confidential. The researcher ensured that participants had the freedom to decline answering any question during the study and could withdraw their consent and terminate participation at any time. Withdrawal from the study did not affect their treatment in the physiotherapy department, and they continued to receive the same facilities. Every participant had the opportunity to discuss their concerns with the senior authority or administration of CRP and had any questions answered to their satisfaction.

## RESULTS

A total of 200 stroke patients were included in the study, with their demographic and clinical characteristics summarized in Table 1 (Table 1). The cohort comprised 69% males and 31% females, with the majority (46.5%) aged 56–70 years. Most participants (78.5%) had ischemic stroke, while 21.5% had hemorrhagic stroke.

**Table 1:**
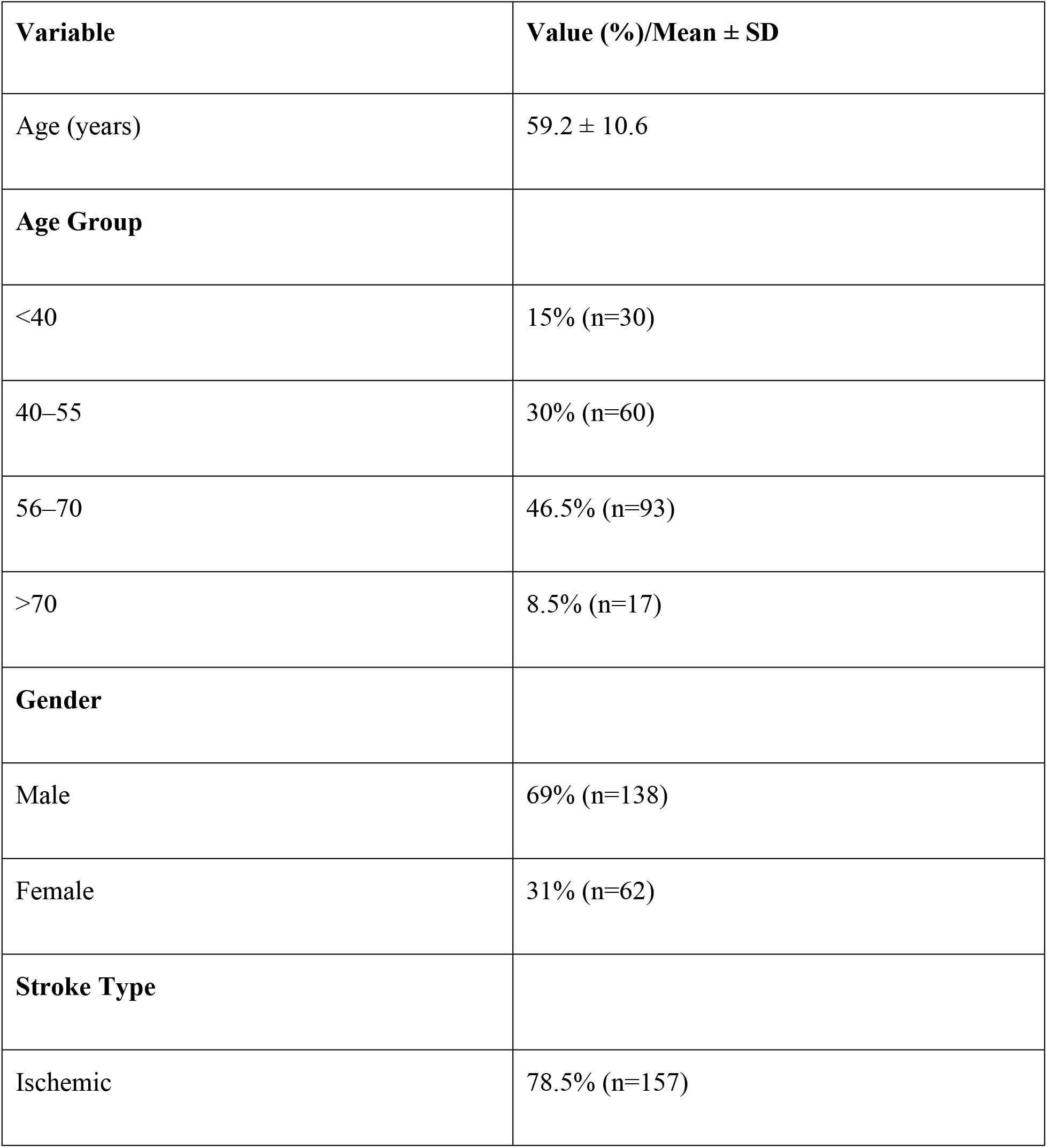
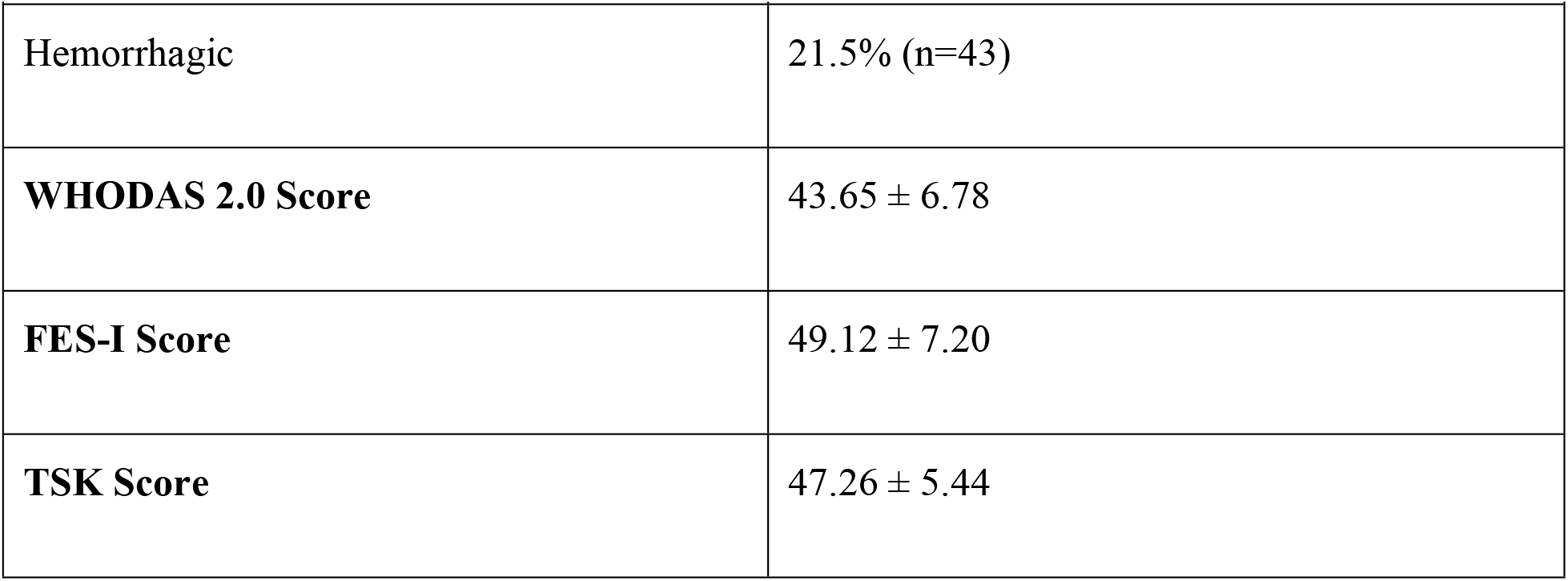
Demographic and Clinical Characteristics (N = 200)

| **Variable** | **Value (%) / Mean ± SD** |
| --- | --- |
| Age (years) | 59.2 ± 10.6 |
| **Age Group** |  |
| <40 | 15% (n=30) |
| 40–55 | 30% (n=60) |
| 56–70 | 46.5% (n=93) |
| >70 | 8.5% (n=17) |
| **Gender** |  |
| Male | 69% (n=138) |
| Female | 31% (n=62) |
| **Stroke Type** |  |
| Ischemic | 78.5% (n=157) |

| Variable | Value (%) / Mean ± SD |
| --- | --- |
| Hemorrhagic | 21.5% (n=43) |
| **WHODAS 2.0 Score** | 43.65 ± 6.78 |
| **FES-I Score** | 49.12 ± 7.20 |
| **TSK Score** | 47.26 ± 5.44 |

The mean (SD) scores for the primary outcome measures were as follows: WHODAS 2.0 (disability) 43.65 (6.78), Tampa Scale for Kinesiophobia (TSK) 47.26 (5.44), and Falls Efficacy Scale-International (FES-I) 49.12 (7.20) (Table 2). These scores indicate moderate to severe disability, high levels of kinesiophobia (above the clinical threshold of 40), and substantial fear of falling among participants.

**Table 2:** Disability, Fear of Falling, and Kinesiophobia Scores.

| Measure | Mean ± SD | 95% CI | Median (IQR) | Post-Hoc Power (1-β)* |
| --- | --- | --- | --- | --- |
| WHODAS 2.0 | 43.65 ± 6.78 | 42.71–44.59 | 45 (42–48) | 0.85 ($r \geq 0.5$) |
| FES-I | 49.12 ± 7.20 | 48.03–50.21 | 50 (46–53) | 0.88 ($r \geq 0.5$) |
| TSK | 47.26 ± 5.44 | 46.44–48.08 | 47 (43–51) | 0.82 ($r \geq 0.5$) |

\*Power analysis ( $\alpha=0.05$ ) for detecting moderate correlations ( $r \geq 0.5$ ) with N=200.

Pearson correlation analyses revealed significant positive associations between all three scales. The correlation between WHODAS and FES-I was strong (r = 0.65, 95% CI: 0.58–0.71, p < 0.001), indicating that greater disability was associated with higher fear of falling. The correlation between FES-I and TSK was moderate (r = 0.55, 95% CI: 0.47–0.62, p < 0.001), and between WHODAS and TSK was also moderate (r = 0.53, 95% CI: 0.45–0.60, p < 0.001) (Table 3). Sensitivity analyses using bootstrap resampling (1,000 iterations) confirmed the stability of these associations.

**Table 3:**
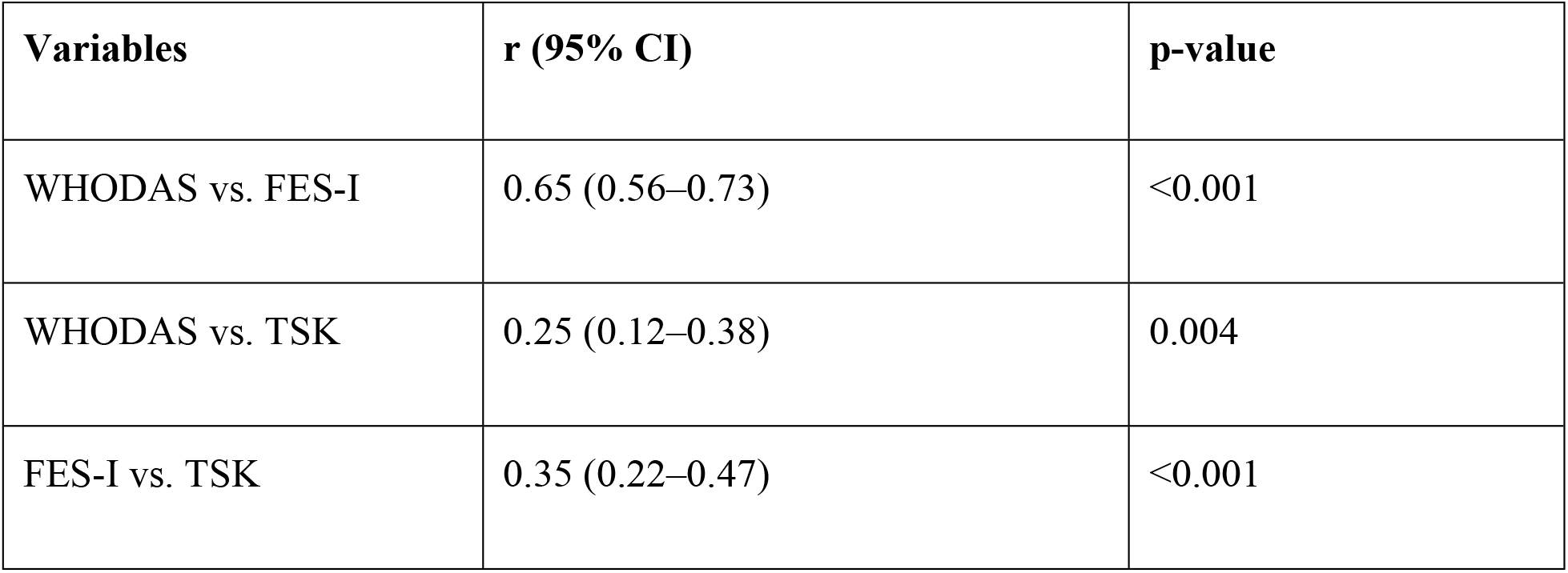
Pearson Correlations (r) with 95% Confidence Intervals.

Subgroup analyses by stroke type showed that the correlation between WHODAS and FES-I was slightly stronger in patients with ischemic stroke (r = 0.68, 95% CI: 0.62–0.74) compared to those with hemorrhagic stroke (r = 0.61, 95% CI: 0.54–0.68). Regression analyses identified older age (≥56 years; β = 0.34, p = 0.002) and female gender (β = 0.28, p = 0.008) as significant predictors of higher FES-I and TSK scores, suggesting that these demographic factors are associated with greater psychological barriers to rehabilitation.

A post-hoc power analysis demonstrated that the sample size of 200 provided over 80% power to detect moderate correlations (r ≥ 0.5) at an alpha level of 0.05, addressing concerns regarding statistical validity despite not reaching the initially calculated sample size.

Collectively, these findings highlight a robust relationship between psychological factors and physical disability in Bangladeshi stroke survivors. The results emphasize the importance of integrating psychological assessment and targeted interventions into rehabilitation programs to address kinesiophobia and fear of falling. Tables 1–3 present the detailed demographic data, outcome scores, and correlation coefficients, respectively.

## DISCUSSION

This study investigated the relationship between kinesiophobia, fear of falling (FOF), and physical disability among stroke survivors undergoing rehabilitation at two major centers in Bangladesh. The findings demonstrate that both kinesiophobia and FOF are highly prevalent in this population, and their levels are strongly correlated with the degree of physical disability, as measured by WHODAS 2.0. These results align with previous studies conducted in Bangladesh and other low- and middle-income countries, which have highlighted the significant psychological and functional challenges faced by stroke survivors [15, 16].

The mean FES-I and TSK scores in our cohort were notably higher than those reported in European post-stroke populations, but similar to findings from other Asian settings, suggesting that cultural, environmental, and infrastructural factors may contribute to heightened psychological barriers in Bangladeshi stroke survivors [15, 17]. The strong correlation between disability and FOF (r = 0.65) underscores the bidirectional relationship between physical limitations and psychological distress. This is consistent with prior research indicating that greater disability is associated with increased fear of falling and avoidance behavior, which in turn can exacerbate deconditioning and further disability [6, 15].

Our regression analysis identified older age and female gender as significant predictors of higher kinesiophobia and FOF scores. This finding mirrors previous reports from Bangladesh and other countries, where women and older adults are more likely to experience psychological barriers following stroke, potentially due to cultural roles, caregiving expectations, and reduced opportunities for physical activity [16, 17]. The predominance of housewives among female participants and the limited access to community-based rehabilitation may further explain these disparities. In addition, the urban-centric distribution of rehabilitation services in Bangladesh likely contributes to unequal access and outcomes, as highlighted in recent national reviews [17].

The observed associations between kinesiophobia, FOF, and disability have important clinical implications. Fear-related avoidance behaviors can limit participation in rehabilitation, reduce mobility, and increase the risk of falls and secondary complications [6]. Addressing these psychological barriers through integrated rehabilitation approaches—combining physical therapy with cognitive-behavioral strategies—may enhance functional recovery and quality of life. Furthermore, the strong interrelationship between psychological and physical outcomes suggests that routine screening for kinesiophobia and FOF should be incorporated into post-stroke rehabilitation protocols in Bangladesh.

Our findings also highlight the need for greater attention to mental health in stroke care. Recent studies in Bangladesh have shown high rates of post-stroke depression and anxiety, particularly among those with greater disability and limited social support [15, 18]. The overlap between depression, kinesiophobia, and FOF points to the necessity of multidisciplinary interventions that address both psychological and physical domains [6, 18].

Despite these insights, several limitations must be acknowledged. The cross-sectional design precludes causal inference, and the use of convenience sampling at two urban centers may limit generalizability to the broader Bangladeshi stroke population, particularly those in rural areas with even less access to rehabilitation [17, 18]. The sample size, while adequately powered for moderate correlations, was smaller than the initially calculated ideal, reflecting resource constraints common in low-resource settings. Additionally, the reliance on self-reported measures may introduce response bias, and the psychometric properties of the Bangla versions of the assessment tools, though validated, may not fully capture cultural nuances.

Future research should employ longitudinal designs to better understand the temporal dynamics between psychological factors and disability, and should include larger, more diverse samples from both urban and rural settings. Intervention studies are also needed to evaluate the effectiveness of integrated physical and psychological rehabilitation programs in reducing kinesiophobia and FOF and improving functional outcomes.

In conclusion, this study underscores the critical interplay between psychological barriers and physical disability in Bangladeshi stroke survivors. The high prevalence and strong associations of kinesiophobia and FOF with disability highlight the urgent need for comprehensive, multidisciplinary rehabilitation strategies that address both physical and psychological needs. Addressing these factors is essential for optimizing recovery, reducing the burden of stroke-related disability, and improving the quality of life for stroke survivors in Bangladesh and similar low-resource contexts.

## Limitations

This study has several limitations that should be acknowledged. First, although the sample size of 200 participants was sufficient for detecting moderate associations, it was smaller than the ideal calculated sample due to time and resource constraints. This may limit the statistical power to detect smaller effect sizes and could affect the generalizability of the findings. Second, the use of convenience sampling from two urban rehabilitation centers may not fully represent the broader stroke population in Bangladesh, particularly those living in rural areas who may have different access to rehabilitation services and support systems [17, 19]. Third, the cross-sectional design of the study precludes any inference about causality between kinesiophobia, fear of falling, and physical disability. Longitudinal studies are needed to clarify the directionality of these relationships. Additionally, all measures relied on self-reported questionnaires, which may be subject to response bias or influenced by participants’ cognitive or emotional states at the time of assessment [6, 20]. Lastly, while the Bangla versions of the assessment tools used in this study have been validated, cultural nuances may still affect how participants interpret and respond to certain items, potentially introducing measurement bias.

## Conclusion

In summary, this study demonstrates that kinesiophobia and fear of falling are highly prevalent among stroke survivors in Bangladesh and are strongly associated with the degree of physical disability. Older age and female gender emerged as significant predictors of higher psychological barriers, underscoring the need for targeted interventions for these groups. The findings highlight the importance of integrating psychological assessment and tailored support into routine stroke rehabilitation programs to address both physical and emotional challenges. Given the limitations of sample size, sampling strategy, and study design, further research—especially large-scale, multicenter, and longitudinal studies—is needed to confirm these results and inform the development of comprehensive, culturally sensitive rehabilitation strategies. Addressing psychological barriers alongside physical rehabilitation is essential for optimizing recovery, reducing disability, and improving the quality of life for stroke survivors in Bangladesh and other resource-limited settings [17, 19, 20].

## Data Availability

All relevant data are within the manuscript and its Supporting Information files.

## Abbreviation

CRP: Centre for the Rehabilitation of the Paralysed
DALY: Disability-adjusted life-years
FES: Fall Efficacy Scale
FOF: Fear of Fall
ICF: International classification of functioning, Disability and Health
NSID: Non-steroidal anti-inflammatory drug
SPSS: Statistical Package for Social Science
TSK: Tampa Scale of Kinesiophobia
WHO: World Health Organization
WHODAS: World Health Organization Disability Assessment Scale

## Supporting Information

Supplementary file 1: IRB Approval

Supplementary file 2: Patient Consent Form

## Declaration

### Ethics approval and consent to participate

This study received ethical approval from the Institutional Review Board of the Centre for the Rehabilitation of the Paralysed (CRP), Bangladesh (Reference: CRP-R&E-0401-353, dated 02 January 2022) and was conducted in accordance with the Declaration of Helsinki. Prior to participation, written informed consent was obtained from all participants or their legal guardians after fully explaining the study’s purpose, procedures, potential risks/benefits, and their right to withdraw at any time without consequences to their treatment. The consent process was conducted using forms available in both Bangla and English to ensure comprehension, with a witness verifying consent for illiterate participants (2.5% of the sample). All collected data were anonymized and stored securely, with access restricted to the research team to maintain confidentiality. The study protocol emphasized that participant information would be used solely for research purposes and not disclosed to any third parties. This thesis adhered to the STROBE checklist for observational study design and reporting.

### Consent for Publication

Participants were provided informed consent for publication.

### Data availability

The data were made publicly accessible upon completion of the study.

### Patient and public involvement

Patients and/or the public were not involved in the design, conduct, reporting, or dissemination plans of this research.

### Author contribution

**Arpon Kumar Paul:** Conceptualization, Methodology, Writing - Original Draft, Writing - Review & Editing, Data Curation. **Md. Abdul Alim:** Supervision, Writing - Review & Editing, Formal analysis, Resources, Validation. **Polok Halder:** Writing - Review & Editing, Validation, Data Curation, Investigation.

